# Fine-mapping reveals four *TMEM106B* haplotypes associated with neurodegenerative risk and preserved cognition at extreme age

**DOI:** 10.1101/2025.02.02.25321330

**Authors:** Alex Salazar, Niccolò Tesi, Lydian Knoop, Natasja M. van Schoor, Martijn Huisman, Betty Tijms, Everard G.B. Vijverberg, Sven van der Lee, Sanduni Wijesekera, Jana Krizova, Mikko Hiltunen, Markus Damme, Leonard Petrucelli, Marcel Reinders, Anke A. Dijkstra, Inge R. Holtman, Marc Hulsman, Henne Holstege

**Author notes:** **CORRESPONDING AUTHOR:** Henne Holstege.

## Abstract

Genetic variation at the *TMEM106B* locus has been associated with Alzheimer’s disease (AD) and related neurodegenerative phenotypes, yet its haplotypic architecture remains incompletely resolved. We refined the genetic landscape of *TMEM106B* by integrating AD-GWAS summary statistics (∼978K individuals) with haplotype analyses in 5,873 Dutch genomes including AD-cases, age-matched controls, and cognitively healthy centenarians. In addition to the known linkage disequilibrium (LD) block defining risk and protective haplotypes characterized by the amino acid substitution at residue 185 (Threonine/Serine), we identify a second conditionally independent LD block. Together these form four haplotypes (T1-T4), with T3 strongly enriched in centenarians. Long-read whole-genome sequencing in 493 individuals reveals haplotype-linked structural variation and methylation patterns, including a ∼19 Kbp genomic rearrangement carried by T3. These findings indicate that the *TMEM106B* locus comprises multiple haplotypes defined by methylation patterns and structural variants, offering insights into GWAS associations with AD risk and preserved cognition at extreme age.

## INTRODUCTION

*TMEM106B* plays an important role in maintaining cognitive health during aging. Genome-wide association studies (GWAS) have repeatedly identified variants at the *TMEM106B* locus that influence risk for multiple neurodegenerative diseases [1–3]. These variants are in complete linkage disequilibrium (LD), forming an LD-block that defines two haplotypes. The risk haplotype has a frequency of ∼59% in European populations, and has been associated with various neurodegenerative diseases, including Alzheimer’s disease [3] and frontotemporal dementia [1,2]. The reciprocal protective haplotype carries the alleles associated with protection against these traits, and occurs at a frequency of ∼41%.

We recently observed that the protective haplotype is among the most enriched AD-associated loci in cognitively healthy centenarians [4]. This enrichment suggests that *TMEM106B* may contribute to preserved cognitive function into extreme old age. Importantly, recent neuropathological analyses indicate that *TMEM106B* variants are primarily associated with TDP-43-related pathologies (including LATE-NC and HS) [5]; tau tangles [5,6]; and aggregation of TMEM106B fibrils [7–9]. This raises the possibility that the enrichment of the protective haplotype in centenarians reflects protection against neurodegeneration mediated by these proteinopathies. Consistent with this, a subset of centenarians from the 100-plus cohort exhibited TDP-43 aggregates in the brain, and TDP-43 positivity was associated with reduced cognitive performance [10].

The specific causal variants underlying the effects in the *TMEM106B* haplotypes remain unresolved [11,12]. One candidate is the missense variant rs3173615 (C>G), which results in a Thr185Ser substitution. The risk-associated rs3173615-C allele encodes threonine at position 185 and has been hypothesized to alter glycosylation, potentially influencing protein stability or aggregation propensity [13]. Indeed, carriers of the risk haplotype exhibit higher levels of TMEM106B C-terminal aggregates compared to carriers of the rs3173615-G allele (185-Ser) [7–9]. However, knock-in of the homologous rs3173615-G allele in mice did not confer protection [12], suggesting that this variant alone may not explain disease risk. The risk haplotype contains multiple additional variants in complete LD with rs3173615-C that could contribute to functional differences. These include an AluYb8 insertion in the 3′ untranslated region (3′ UTR) [14–16] and a variant affecting CTCF binding near rs1990620 [17], both of which may influence transcriptional or post-transcriptional regulation of *TMEM106B*.

Despite consistent GWAS associations, the genetic architecture underlying the *TMEM106B* locus remains incompletely resolved. In particular, variants within the locus are typically inherited in strong linkage disequilibrium, complicating efforts to disentangle independent signals and identify functional alleles. Moreover, structural variants (SVs), including genomic rearrangements, tandem repeats, and transposable elements, are largely invisible to conventional imputation-based approaches and may contribute to haplotype-specific effects that remain unexplored.

Here, we refine the *TMEM106B* locus by integrating long-read sequencing and large-scale GWAS summary statistics with haplotype analyses in cognitively healthy centenarians, AD cases, and age-matched controls.

## RESULTS

### Two independent LD blocks in the TMEM106B locus are associated with Alzheimer’s disease (AD)

To identify additional AD-associated haplotypes beyond the known risk and protective haplotypes, we integrated linkage disequilibrium (LD) profiles from 5,873 Dutch genomes, comprising cognitively healthy centenarians, AD-cases, and age-matched controls, with summary statistics from the latest AD GWAS [22] (∼978,000 participants; **Figure 1a**). We restricted analyses to common variants that: (i) surpassed genome-wide significance in their associations with AD (p≤5.5×10^−8^), (ii) had minor allele frequency ≥1%, and (iii) clustered within multi-SNP LD blocks defined by r^2^≥0.15 (see **Methods**).

**Figure 1.**
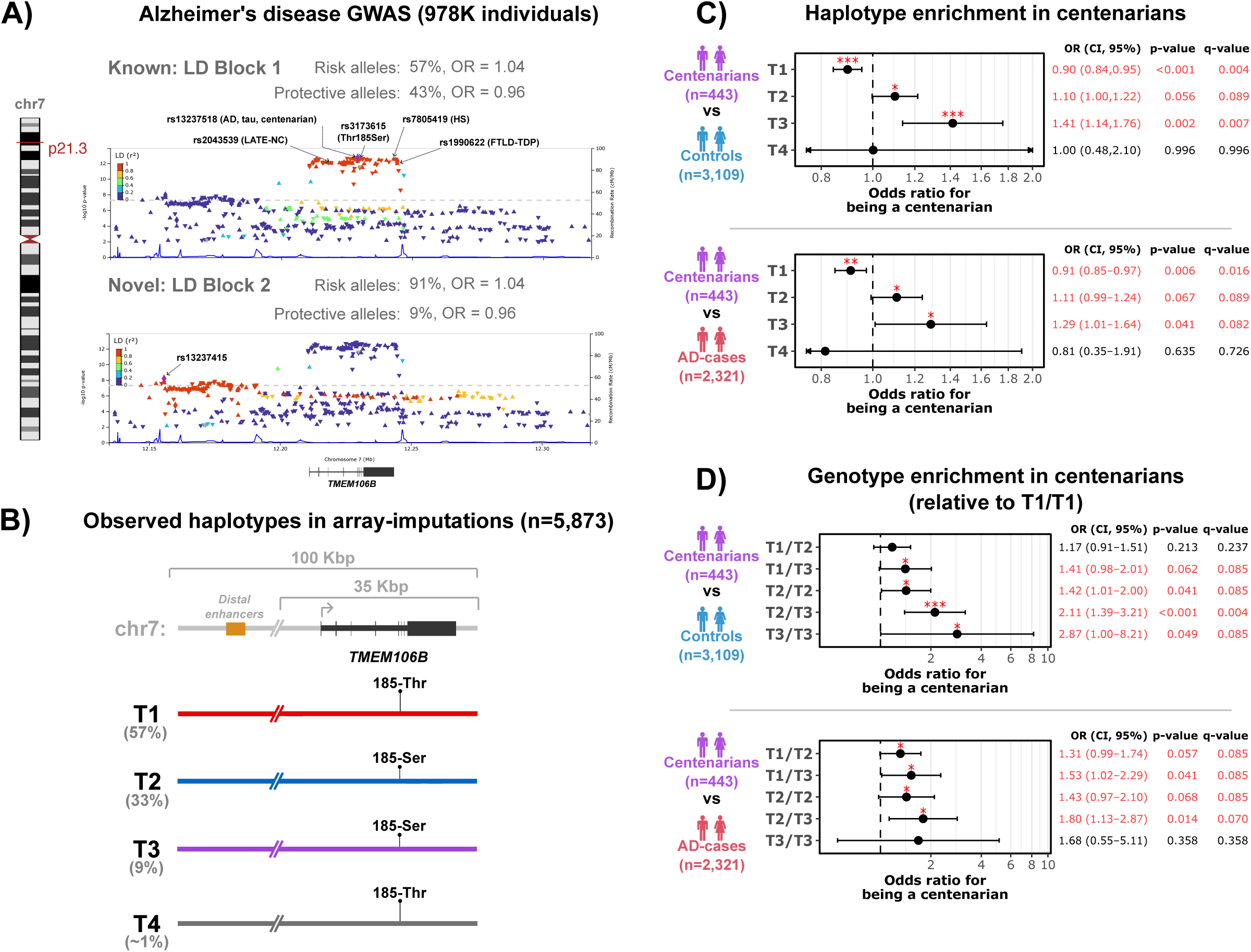
Four separate *TMEM106B* haplotypes are differentially associated with risk for Alzheimer’s disease (AD) and enrichment among cognitively healthy centenarians. **(A)** LocusZoom plot of the *TMEM106B* locus in the latest AD-GWAS [22] highlighting conditionally independent LD blocks. Top: Block 1 encompassing the known risk and protective haplotypes which harbor alleles previously associated with risk to various neurodegenerative traits and pathologies, marked by rs5011439. Bottom: Block 2 encompassing novel risk and protective haplotypes marked by rs13237415 with similar effects as Block 1. **(B)** The alleles from both blocks are inherited in four specific haplotypes, T1-T4, as observed in 5,873 Dutch genomes. **(C-D)** Enrichment analysis of the haplotypes when comparing centenarians with controls or AD-cases. FDR-adjusted q-value (10% threshold): *** < 0.01; ** < 0.05; * < 0.1. **(C)** At the individual haplotype level, T3 is significantly enriched in centenarians, while T1 is significantly depleted. **(D)** Relative to T1/T1 homozygous carriers, the T2/T3 genotype is significantly enriched in centenarians.

Two independent LD blocks at *TMEM106B* reached genome-wide significance (**Figure 1a**). Block 1 (marker SNP rs5011439-C; AF=43%, OR=0.96, p=1.9×10^−13^) overlaps the *TMEM106B* coding region and corresponds to known risk and protective haplotypes (risk: rs5011439-G; protective: rs5011439-C). Block 2, marked by SNP rs13237415-T (AF=9%, OR=0.94, p=7.7×10^−9^), spans ∼65 Kbp upstream to ∼25 Kbp downstream of *TMEM106B* (**Figure 1a**). The blocks are largely independent (r^2^=0.11), and each remained significant in joint/conditional GCTA-COJO analyses (OR=0.96 for both; p=6.4×10^−9^ and 3.3×10^−4^), even when extending to other marker SNPs in their respective LD blocks (**Methods**). All SNPs in Block 2 are in strong LD (r^2^≥0.8), defining two additional haplotypes with rs13237415-G associated with risk and rs13237415-T with protection.

These findings thus identify two conditionally independent association signals at the *TMEM106B* locus, each defined by distinct haplotypes.

### Four TMEM106B haplotypes are differentially enriched in cognitively healthy centenarians

The allele combination of the sentinel SNPs of Blocks 1 and 2 defines four haplotypes across all genomes (n=11,746; **Figure 1b, Supplementary Table S1**). We refer to these haplotypes as T1-T4 in descending frequency: T1 is the most common combination (57%, n=6,701) and contains the risk-associated alleles of both blocks. T2 follows (33%, n=3,919) and carries the protective alleles of block 1 and risk alleles of block 2. T3 (9%, n=1,020) carries protective alleles from both blocks, and T4 (∼1%, n=106) carries risk alleles from block 1 and protective alleles from block 2.

We found that, compared with non-demented controls, carriers of T3 were significantly associated with a 1.41-fold higher odds of being a centenarian (OR=1.41, 95% CI=[1.14,1.76]; p=0.002; q=0.007). In contrast, T1 was significantly depleted in centenarians, with a 0.9-fold lower odds (OR=0.90, CI=[0.84,0.95]; p=5.61×10^−4^; q=0.004). T2 had modest but significant enrichment (OR=1.10, CI=[1.00,1.22]; p=0.056; q=0.089), while enrichment for T4 was imprecise due to its relatively low frequency (OR=1.00, CI=[0.48,2.10]; p=0.996; q=0.996).

To further disentangle the contribution of the 185-Serine-encoding haplotypes, we repeated the analysis using T2, the most frequent serine-containing haplotype, as the reference haplotype. Under this model, T1 remained depleted (OR=0.81, CI=[0.70,0.95]; p=0.009; q=0.036), while T3 remained enriched (OR=1.29, CI=[1.00,1.66]; p=0.052; q=0.070), indicating that the enrichment of T3 cannot be explained by the serine allele alone and may reflect additional protective features. Similar patterns were observed when comparing centenarians with AD cases, although effect sizes were attenuated **(Figure 1c)**.

Analogous to the genotype-specific effects observed for ε4/ε4, ε4/ε3, and ε4/ε2 at *APOE*, we next examined the *TMEM106B* genotypes in centenarians. To do this, we fitted a logistic regression model relative to T1/T1 homozygous carriers, the genotype with the most depleted haplotype. This analysis reinforced the prominent role of T3 (**Figure 1d**): the T2/T3 genotype was associated with a 2.11-fold higher odds of being a centenarian (OR=2.11, CI=[1.39,3.21]; p=4.38×10^−4^; q=0.004), and the estimate was larger for T3/T3 homozygotes (OR=2.87, CI=[1.00,8.21]; p=0.049; q=0.085), although the confidence interval was wide. Other genotypes, including T1/T2, T1/T3, and T2/T2, showed more modest enrichments (**Figure 1d**). In contrast, estimates for genotypes containing T4 were imprecise due to its low frequency. Similar patterns were observed when comparing centenarians with AD cases, although effect sizes were attenuated (**Figure 1d**).

### Long-read fine-mapping reveals haplotype-linked structural variants in TMEM106B

To fine-map the genetic basis of the differential associations observed for the T1-T4 *TMEM106B* haplotypes, we performed long-read whole-genome sequencing of 493 individuals, including 248 cognitively healthy centenarians from the 100-plus Study [18] and 245 AD-cases from the Amsterdam Dementia Cohort [19] (**Methods**). We used these data to characterize haplotype-resolved structural variation and DNA methylation patterns of the different haplotypes spanning *TMEM106B*.

We found eight common structural variants (SVs, ≥25 bp, ≥1% frequency). The SVs ranged from 35 bp to ∼19 Kbp (**Figure 2a**), with allele frequencies spanning ∼10-90%, similar to those of the two LD blocks (**Supplementary Table S2**). In addition to the previously reported AluYb8 insertion in the 3′UTR, we identified seven novel SVs. The largest was a ∼19 Kbp genomic rearrangement located ∼51 Kbp upstream of the *TMEM106B* transcription start site, likely mediated by an AluYh3 element (**Figure 2b**). Among the identified SVs, a multi-allelic tandem repeat (TR_12157495) located near the rearrangement directly overlapped a known distal enhancer (EH38E2534480), suggesting potential regulatory relevance.

**Figure 2.**
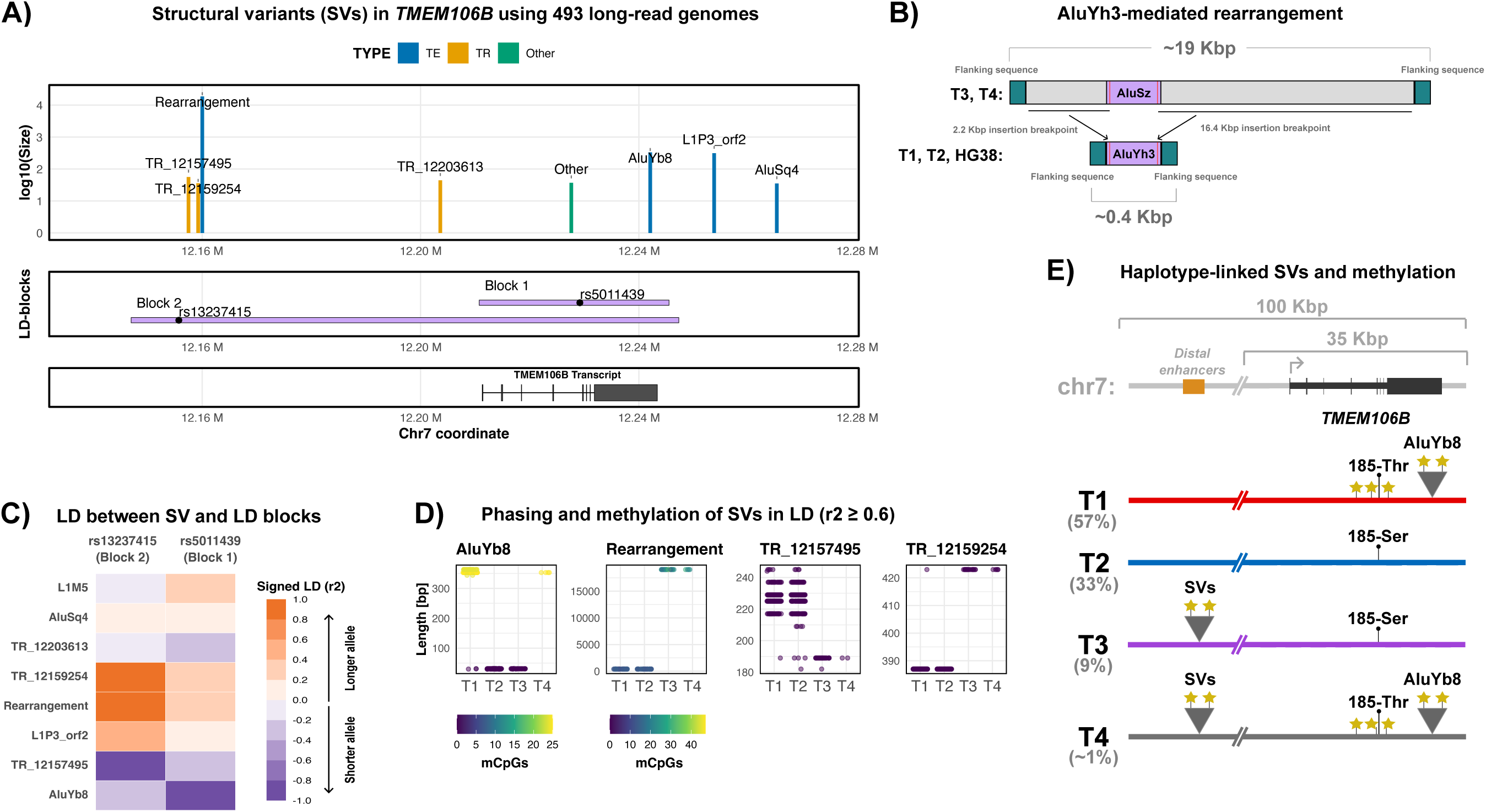
The *TMEM106B* haplotypes are characterized by large structural variants (SVs). **(A)** Landscape of SVs in the *TMEM106B* locus using 493 long-read sequenced genomes. **(B)** The largest SV was a ∼19 Kbp genomic rearrangement likely mediated by an AluYh3 element. **(C)** LD values (r^2^) between the SVs and the SNP markers for the Block 1 and Block 2. **(D)** Long-read phasing and CpG methylation of allele sequences for SVs in moderate LD (r^2^≥0.6) with either Block 1 or Block 2. **(E)** These analyses highlight that SVs distinctly characterize the T1-T4 haplotypes of *TMEM106B*.

Both LD-based analyses and long-read-based phasing independently supported four haplotype-linked SVs (**Figure 2c,d**): the AluYb8 insertion was linked to T1 and T4; and the ∼19 Kbp rearrangement, along with contractions in TR_12157495 and expansions in TR_12159254, were linked to T3 and T4 (**Figure 2c**). The AluYb8 insertion and the ∼19 Kbp rearrangement were both highly methylated (**Figure 2d**), highlighting possible regulatory effects.

We validated the T1-T4 haplotype architectures and their linked SVs using complementary approaches, including long-read whole-genome *de novo* assembly of the 493 genomes (**Supplementary Results**). Haplotype genotype calls (for example, T1/T1 and T1/T2) showed ∼99% concordance across approaches, and phased SV-haplotype assignments showed ≥93% concordance (**Supplementary Results**). Consistent with this, read-based phasing and fully phased *de novo* assemblies spanning the *TMEM106B* locus, available for 338 of 493 genomes, yielded nearly identical combinations of SNP and SV alleles for the common T1-T4 haplotypes (**Figure 3b,c**).

**Figure 3.**
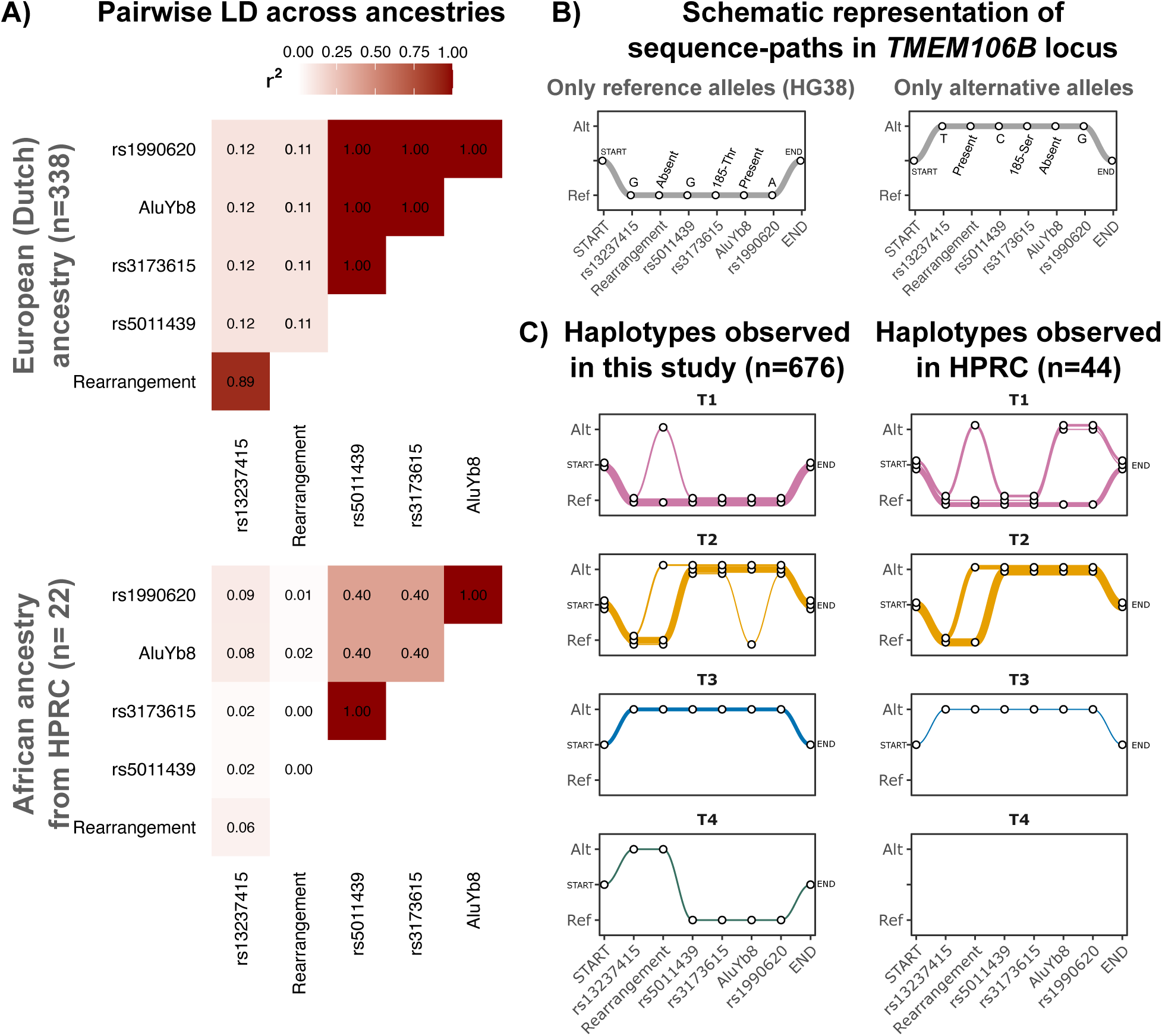
Distinct *TMEM106B* haplotypes are observed in genomes of European and African ancestry. **(A)** Pairwise LD of several notable variants from Figure 1a and Figure 2a based on 338 available *de novo* assemblies from the 493 Dutch long-read sequenced genomes, and 22 long-read *de novo* assemblies from the Human Pangenome Research Consortium (HPRC) [35]. **(B)** Schematic representation of allele composition within individual haplotypes illustrated as sequence-paths. Left: sequence-path containing only reference alleles, corresponding to the GRCh38 reference genome. Right: sequence path containing only alternative alleles. **(C)** Left: observed sequence-paths for the *TMEM106B* haplotypes (T1-T4) in the 338 *de novo* assemblies (European-Dutch ancestry). Right: sequence-paths observed in 22 genomes in HPRC.

Closer inspection of the fully phased assemblies identified several rare configurations that differed from the common T1 and T2 architectures. Three T1-like and three T2-like haplotypes carried the ∼19 Kbp rearrangement, whereas one additional T2-like haplotype carried the AluYb8 insertion (**Figure 3b,c**). These configurations may represent rare *TMEM106B* haplotypes in the Dutch population, and/or assembly errors.

Prior work has identified differences in LD structure at the *TMEM106B* locus between genomes of European and African ancestry [32]. We therefore compared the T1-T4 haplotypes derived from the Dutch de novo assemblies with those observed in 22 long-read-based assemblies from individuals of African ancestry from the Human Pangenome Research Consortium (HPRC) [35]. Consistent with previous observations, LD between the major variant groups was reduced in the HPRC genomes (**Figure 3a; Supplementary Results**). The rare T2-like configuration carrying the ∼19 Kbp rearrangement observed in the Dutch genomes was also present among the HPRC haplotypes (**Figure 3b,c**). T1-like haplotypes carrying the ∼19 Kbp rearrangement were also observed, but these additionally lacked the AluYb8 insertion and carried the protection-associated rs1990620 allele (**Figure 3b,c; Supplementary Results**). Thus, these haplotypes combine the risk-associated 185-threonine allele with the rs1990620 allele and absence of the AluYb8 element, both of which are associated with protection. Larger datasets will be required to determine the associations of these distinct haplotypes with neurodegenerative disease; however, these findings indicate that *TMEM106B* haplotype architecture and frequencies can vary across populations.

Although T2 and T3 both carry the 185-serine allele, T3 is distinguished by multiple linked SVs located ∼51 Kbp upstream of *TMEM106B* (**Figure 2e**). These haplotype-linked SVs may contribute to the preferential enrichment of T3 in centenarians. Functional studies will be required to determine their biological consequences.

### The TMEM106B haplotypes have distinct methylation profiles

We next investigated whether *TMEM106B* haplotypes harbor distinct DNA methylation profiles using CpG methylation frequencies derived from haplotype-resolved long-read sequencing data. Pairwise comparisons of the common haplotypes (T1, T2, and T3) identified 146 unique CpG sites that were differentially methylated in at least one comparison (median methylation difference ≥10%), of which 129 showed a median difference of ≥90% (**Figure 4a and Supplementary Table S3**). Many of these differences were driven by SNPs that created or disrupted CpG dinucleotides, indicating haplotype-specific CpG content (**Supplementary Table S3**). These sites included CpGs overlapping annotated distal enhancers: two associated with T1, three with T2 and four with T3. T1 showed higher methylation across introns 3 and 4 and the 3′ UTR, including the AluYb8 element, whereas T3 showed lower methylation near the upstream rearrangement (**Figure 4a**).

**Figure 4.**
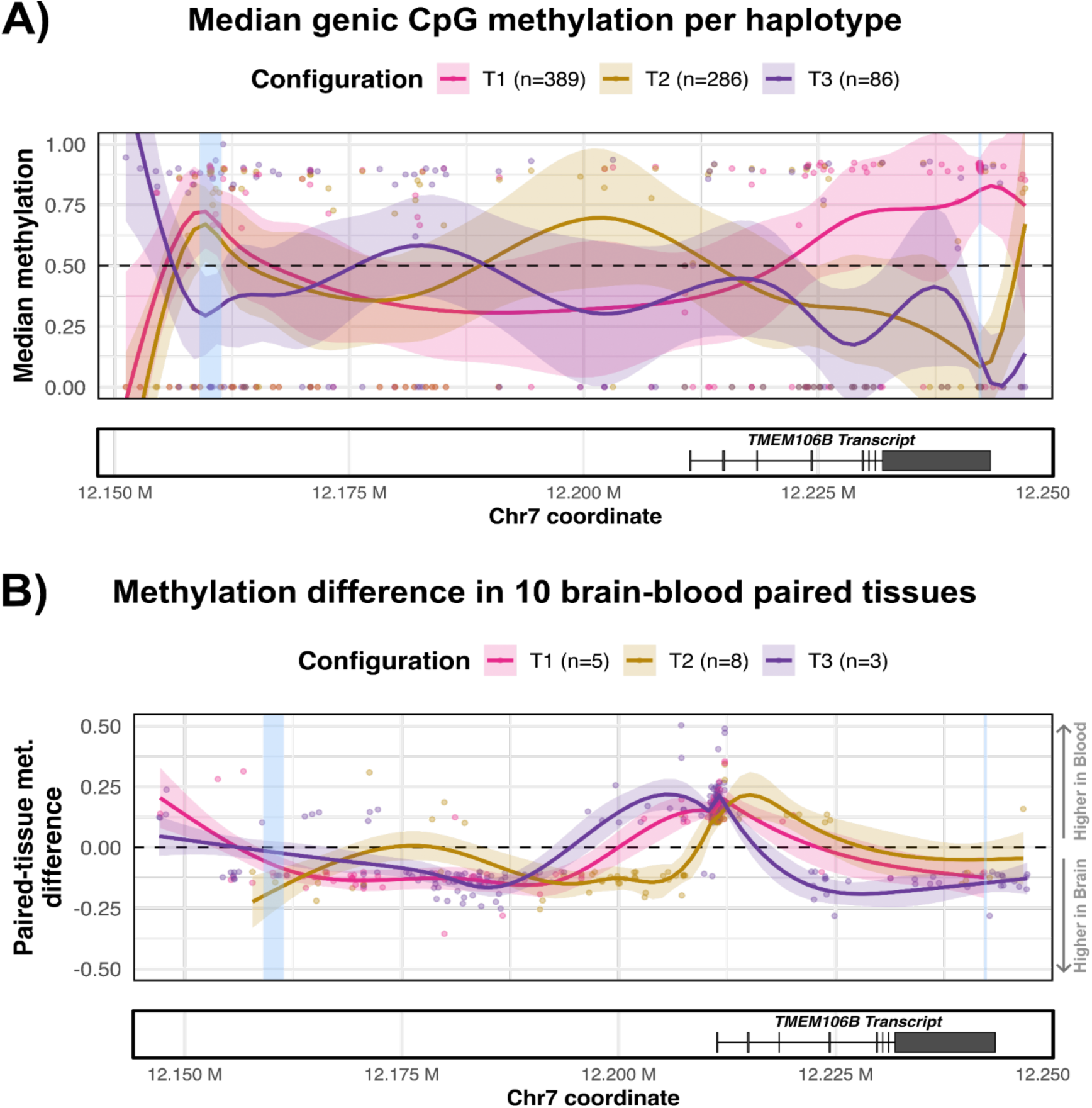
The *TMEM106B* haplotypes are also characterized by distinct CpG DNA methylation profiles. **(A-B)** CpG methylation profiles derived from phased long-read sequencing of blood and brain tissue samples. Differentially methylated CpG sites are defined as sites where the fraction of methylated reads is at least 10% between haplotypes or paired tissues. **(A)** CpG methylation profiles derived from blood samples of 493 individuals. Dots show median methylation levels at 146 differentially methylated CpG sites across the *TMEM106B* locus for the three common haplotypes (T1-T3). Curves represent spline fits, and shaded bands indicate standard errors. The lower track shows the canonical *TMEM106B* transcript. **(B)** For a subset of 10 individuals, long-read whole-genome sequencing was also performed on matched brain tissue (temporal cortex). Shown are per-sample methylation differences between brain and blood for CpG sites exhibiting an absolute methylation difference ≥10% (n=301), stratified by haplotype.

As the long-read sequencing data were derived from blood, we examined whether similar methylation patterns were observed in temporal cortex tissue by performing long-read sequencing on paired brain samples from 10 centenarians (**Figure 4b**). Across 16 distinct haplotypes, we identified 301 unique CpG sites that were differentially methylated between brain and blood (absolute median methylation difference ≥10%). Tissue-specific patterns were consistent across haplotypes: methylation levels were higher in the brain at the transcription start site, whereas blood showed higher methylation across introns 3 through the 3′UTR. These results indicate that haplotype-associated methylation patterns in *TMEM106B* likely differ between tissues.

## DISCUSSION

In this study, we refine the genetic architecture of the *TMEM106B* locus and demonstrate that two conditionally independent GWAS LD blocks generate four common haplotypes (T1-T4) with differential associations with AD and differential enrichment among cognitively healthy centenarians. Among these haplotypes, T3 shows the strongest enrichment in centenarians, while T1 is depleted. These haplotypes are further distinguished by large structural variants (SVs) and distinct haplotype-associated CpG methylation profiles. These features include a ∼19 Kbp genomic rearrangement, strongly linked to T3 and T4, located ∼51 Kbp upstream of the *TMEM106B* transcription start site near annotated distal enhancers. In addition, T1 is characterized by highly methylated CpG sites within the intronic region and 3′UTR of *TMEM106B*. Together, these results refine the haplotypic architecture underlying *TMEM106B*-associated risk beyond the previously identified GWAS signals.

Our results do not pinpoint the exact causal variants underlying the associations at the *TMEM106B* locus, but instead indicate that multiple genetic mechanisms likely contribute beyond the previously emphasized amino acid substitution at residue 185 (Threonine/Serine). In particular, the role of the Serine allele and its protective associations with neurodegenerative traits has been actively debated [11,12]. Although both T2 and T3 harbor the serine allele, T3 is more strongly enriched than T2 in centenarians, suggesting that additional protective mechanisms may be encoded within the T3 haplotype. The large structural variants strongly linked to T3 represent plausible candidates for such effects; however, we note that multiple other variants, including CpG sites in regulatory elements, are also in strong linkage disequilibrium with T3 and may contribute to the observed associations.

*TMEM106B* has been implicated in TMEM106B fibril pathology, TDP-43-related neurodegeneration, and tau pathology [5–9]. The presence of multiple *TMEM106B* haplotypes raises the possibility that they may differentially influence specific pathological pathways, or exert overlapping effects across these processes. The strong enrichment of T3 in cognitively healthy centenarians suggests that this haplotype may contribute to preserving cognitive function [33,34]. Whether T3 is differentially associated with specific proteinopathies remains to be determined. Testing pathology-specific effects will require cohorts with detailed neuropathological characterization.

Our analyses have three main limitations. First, our structural variant analysis focused on common SVs (allele frequency ≥1%), and mainly in genomes with European ancestry. Rarer variants, and non-European haplotypes may also contribute to locus heterogeneity [32]. Indeed, we observe additional haplotypes with distinct permutations of the 185-Threonine/Serine amino acids along with the SVs including the AluYb8 element and rearrangement. Larger and more diverse cohorts will be needed to estimate the disease associations of these distinct haplotypes. Second, the T4 haplotype is relatively rare, limiting power to estimate its independent effects with precision. Third, although we identify haplotype-linked structural and methylation differences, functional consequences remain to be experimentally validated. The regulatory impact of the upstream rearrangement, tandem repeats, and AluYb8 elements will require targeted functional assays.

Overall, our findings provide new insight into the genetic basis of *TMEM106B*-associated risk and cognitive resilience. By leveraging extreme longevity as an informative phenotype and combining haplotype-aware GWAS interpretation with long-read sequencing, we uncover structural and epigenetic features that refine the locus beyond single-marker associations. These results underscore the importance of haplotype-resolved analyses and structural variation in interpreting complex disease loci and suggest that similar hidden architectures may underlie other GWAS signals in neurodegeneration.

## METHODS

### Study population

Samples used in this study included a total of 5,873 genomes: 443 centenarians from the 100-plus Study [18]; 2,321 Alzheimer disease (AD) patients from the Amsterdam Dementia Cohort [19] and the Netherlands Brain Bank [20]; and 3,109 non-demented controls age-matched with the AD patients previously described [4], including 1,614 from the LASA cohort [21]. The Medical Ethics Committee of the Amsterdam University Medical Center approved all studies. All participants and/or their legal representatives provided written informed consent for participation in clinical and genetic studies.

### Genotyping and AD-GWAS summary statistics

Array-based genotyping and haplotype-imputations of the above cohorts were previously described [4].

We analyzed GWAS variants within 100 Kbp upstream and downstream of *TMEM106B* coding sequence (chr7:12111294-12343367), using the latest AD-GWAS summary statistics [22]. Pairwise linkage disequilibrium (LD) values (r^2^) were calculated with PLINK (v2.00a6LM) [23] using the centenarian, AD-cases, and control cohorts described above, and variants were clumped using a conservative r^2^ threshold of 0.15. We retained the sentinel SNPs of LD blocks with genome-wide significance in the AD-GWAS (p-value < 5.5×10^−8^). We define an LD block as a set of variants that were in strong LD (r^2^≥0.8) with the sentinel SNPs, resulting in two blocks: Block 1 (marked by rs5011439) and Block 2 (rs13237415). To avoid overestimation in downstream analysis, we additionally performed pairwise LD between all variants within each LD block, and identified a representative SNP for each block via hierarchical clustering, resulting in two medoid-based marker SNPs for Block 1 (rs12699338) and Block 2 (rs12699361). Joint effect analyses using GCTA-COJO (v1.94.1) [24] were performed separately for (i) sentinel SNPs from each block; and (ii) the corresponding medoid SNPs, confirming that both sets show independent associations.

### Differential enrichment of TMEM106B haplotype in centenarians

Using the array-imputed haplotypes of the 5,873 genomes (11,746 total haplotypes), we used the sentinel SNPs to classify *TMEM106B* haplotypes (T1-T4), as detailed in **Supplementary Table S1**.

To determine whether individual haplotypes and their genotypes were enriched in centenarians, we used logistic regression models implemented in R (v4.5.2) with the *glm* function by separately comparing centenarians with non-demented controls and AD-cases. All models were adjusted for population structure using the first five genetic principal components calculated with PLINK. As centenarians are known to be enriched for the *APOE* ε2 allele and depleted of ε4 allele [4], we additionally adjusted for dosage of ε2 and ε4 alleles in order to obtain estimates after accounting for *APOE*.

For the enrichment analysis of individual haplotypes, we used a weighted sum-to-zero (effect) encoded joint model to test the independent associations of the T1-T4 haplotypes. This approach enables estimation of the independent effect of each haplotype relative to the frequency-weighted average effect across all haplotypes, allowing effect sizes (β), standard errors, 95% confidence intervals, and p-values to be interpreted on a common scale. Briefly, we fitted a joint logistic regression model with centenarian phenotype as a binary response variable, *Centenarian ∼ T1 + T3 + T4 + ε4_dosage_ + ε2_dosage_ + PC_1-5_*, using T2 as the reference haplotype. We then used the estimated β’s and their variance-covariance matrix to compute the frequency-weighted average effect across all haplotypes, including T2, where weights were defined by haplotype frequencies in 5,873 array-imputed genomes. We re-expressed the haplotype-specific β’s under a weighted sum-to-zero constraint, calculated odds ratios and 95% confidence intervals, and applied Wald tests to evaluate whether each haplotype significantly deviated from the weighted average effect. Coefficients from the original joint model with T2 as the reference were used for the sensitivity analysis to evaluate whether the associations of T1 and T3 were independent of T2.

To determine specific genotypes of the *TMEM106B* haplotypes, we constructed a categorical variable representing all observed genotypes (e.g., T1/T1, T1/T2, T2/T3, etc.). We then fitted a joint logistic regression model using T1/T1 as the reference genotype, similarly adjusting for *APOE* ε2 and *ε4* dosages and genetic PC_1-5_. Odds ratios and 95% confidence intervals were calculated from the regression coefficients.

Because of its relatively low frequency, T4 and T4-containing genotypes were retained in all models, but their estimates were not interpreted.

To account for multiple testing, we defined two families of tests: (i) enrichment of individual haplotypes; and (ii) enrichment of genotypes. We pooled all p-values within each family across both comparisons (centenarians vs non-demented controls; and centenarians vs AD-cases), and corrected them jointly using the Benjamini-Hochberg procedure to control for false discovery rate (FDR). As the *TMEM106B* locus was previously shown to be differentially enriched in centenarians [4] and differentially associated with AD-status [22], we considered FDR-adjusted q-values <0.1 statistically significant. Adjusted q-values are reported alongside nominal p-values.

### Long-read sequencing of 493 Dutch genomes

Peripheral blood tissue of a subset of 248 centenarians and 245 AD-cases were long-read whole-genome sequenced on Pacific Biosciences (PacBio) Sequel II and IIe System. Temporal-cortex tissue from 10 centenarians was sequenced using the same approach. All samples were sequenced at the Department of Clinical Genetics at the Amsterdam University Medical Center (AUMC) and at the Radboud Technology Genome Center and Radboud University Medical Center (RUMC), using 2 hours of pre-extension time and 30 hours of collection time. Overall, the AD and centenarian genomes were sequenced at a median coverage of 18.2x, with median read-lengths of 14.8 Kbp. See extended details in **Supplementary Methods**.

After sequencing, raw reads were collected and analyzed through an in-house pipeline (freely available at https://github.com/holstegelab/snakemake_pipeline; **Supplementary Methods**).

Additionally, we used 47 publicly available long-read whole-genomes from the Human Pangenome Research Consortium [35]. We downloaded publicly available assemblies from “Year 1” data freeze, and GRCh38-aligned BAM-files (https://github.com/human-pangenomics/HPP_Year1_Data_Freeze_v1.0). Long-reads were extracted from the BAM-files using *samtools* (v1.13) [36] and re-aligned to GRCh38 using *minimap2* [37] (version 2.21-r1071). *De novo* assemblies were aligned to GRCh38 using *minimap2* with the ‘-x asm10’ parameter.

### Variant and methylation profiles using long-read sequencing data

Single nucleotide polymorphisms (SNPs) were identified in both the AD-cases and centenarian long-read sequencing data using *DeepVariant* (v1.1.0) [25], and joint-called using *GLnexus* (v1.4.1-0-g68e25e5) using ‘--config Deepvariant’ parameter [26].

Structural variants (SVs) in AD cases, centenarians and HPRC genomes were identified and genotyped using an in-house local-assembly workflow based on *Sniffles2* (v2.0.6) [27] and *otter* (v1.0) [28] **(Supplementary Methods).** To support consistent analysis across sequencing depths, including low-coverage long-read genomes, we developed *haplr* (v0.1.0), a haplotype-guided framework that probabilistically assigns individual long reads to predefined haplotypes and supports haplotype-resolved SV genotyping and CpG methylation profiling (**Supplementary Methods**; https://github.com/holstegelab/haplr).

The above SV workflow comprised three steps. First, allele sequences were locally assembled for each candidate SV in each genome using *otter*. Second, SV alleles were jointly genotyped using a sequence-based clustering procedure implemented in *otter*, enabling allele sequences to be compared and harmonized across more than one million candidate SV regions and large collections of long-read genomes. This step generated a joint SV gVCF for the centenarian and AD genomes. Third, SV genotypes were recalibrated using the *svg* function in *haplr*. Briefly, for each candidate SV, supporting long-reads were locally aligned to the all possible allele sequences in the gVCF, and the most likely genotype and its corresponding genotype quality score were inferred probabilistically. This procedure enabled consistent genotyping across high- and low-coverage samples, and additionally quantified CpG methylation within each SV allele **(Supplementary Methods).**

Pairwise linkage disequilibrium (LD) between SV-SV and SV-SNP genotypes was calculated using length-derived genotype scores. For each SV in the gVCF file, we first identified the minimum and maximum allele lengths among alleles with a frequency ≥1%. Alleles shorter or longer than these bounds were assigned the minimum or maximum length, respectively, thereby limiting the influence of rare, extreme-length alleles. The resulting allele lengths were then scaled between 0 and 1 using these minimum and maximum values. For each genome, the SV genotype score was calculated as the sum of the scaled lengths of its two alleles, yielding a value between 0 and 2. Pairwise LD between SV genotype scores, or between SV genotype scores and SNP allele dosages, was quantified as the squared Pearson correlation coefficient (r^2^). This approach is analogous to the unphased dosage-based r^2^ statistic implemented in PLINK [23], but uses allele length as a quantitative representation of SV allelic state. This procedure is implemented in *haplr*.

The corresponding allele sequences in each SV were annotated using DFAM (release 3.7) [29], including transposable elements (TEs), while tandem repeats (TRs) were annotated by overlapping TR-annotation tracks from the UCSC Genome Browser (http://genome.ucsc.edu) [30].

We used *haplr* to generate haplotype-resolved SV and methylation profiles. Briefly, the *assign* command within *haplr* probabilistically assigns a source haplotype (T1-T4) to each long-read alignment within each sample **(Supplementary Methods)**. The *svg* command uses the haplotype-assigned reads to infer the most likely haplotype for an SV allele. Similarly, these assigned reads are also used to generate haplotype-specific CpG methylation profiles for observed methylated bases using the *mtb* command **(Supplementary Methods).** As an independent validation, assignments made by *haplr* were compared with haplotypes resolved from whole-genome *de novo* assemblies using *hifiasm* [38] (v0.16.1-r375), and reference-based long-read phasing using *HiPhase* [39] (v1.4.2-c7e0700), using the 493 long-read sequenced genomes **(Supplementary Results and Methods)**.

We analyzed long-read *de novo* assemblies to compare T1-T4 haplotypes in genomes with African ancestry. Briefly, we used completely phased *de novo* assemblies from the 493 long-read sequenced genomes (see above), as well as 22 long-read *de novo* assemblies of genomes with African ancestry from the Human Pangenome Research Consortium [35] (**Supplementary Results**), and aligned contigs to the GRCh38 reference genome using *minimap2* with the ‘-x asm10’ parameter. We then used the *wgat* command from *haplr*, which extracts the corresponding allele for a given SV per aligned contig (i.e. phased haplotype), and compares the sequence to all possible alleles observed for that SV in a given gVCF file via local sequence alignment (**Supplementary Methods**). The allele in the gVCF file with the minimum edit distance is then chosen as the corresponding allele index in the aligned contig. We used the joint gVCF generated above to obtain harmonized SV calls for all assemblies, and calculated LD between SVs and the GWAS sentinel SNPs as defined in **Supplementary Table S1** using the same scaled SV allele-length procedure described above.

For methylation analyses, we focused on CpG sites within the start/end coordinates from the union of both LD blocks (chr7:12146976-12247317). CpG sites were retained if they had a minimum per-sample coverage of 4 reads and were observed in at least 10 independent individuals. Methylation levels were calculated per individual and haplotype as the fraction of methylated reads over total reads. For CpG sites absent from one haplotype because of sequence variation that disrupted the CpG dinucleotide, the methylation value for that haplotype was set to 0 for computational analysis. These values represent absence of the CpG site rather than an observed unmethylated state. Accordingly, sequence-dependent differences in CpG content were distinguished from methylation differences at CpG sites shared across haplotypes. CpG methylation was summarised separately for each pairwise haplotype comparison (T1 vs T2; T1 vs T3; and T2 vs T3) across individuals, and sites with at least 10% median methylation difference between haplotypes (|Δ methylation| ≥ 0.1) were considered differentially methylated CpG sites, and those with a 90% difference (|Δ methylation| ≥ 0.9) were considered haplotype-specific. Regulatory annotation was incorporated using candidate cis-regulatory element (cCRE) tracks from ENCODE (dataset ENCFF924IMH) [31].

## DATA AVAILABILITY

Long-read sequencing data generated with PacBio Sequel IIe for the AD patients and cognitively healthy centenarians are available upon reasonable request through the Alzheimer Genetics Hub (AGH, https://alzheimergenetics.org/). Long-read sequencing data from the Human Pangenome Research Consortium is publicly available (https://github.com/human-pange-nomics/HPP_Year1_Data_Freeze_v1.0).

## CODE AVAILABILITY

Custom software used in this study is publicly available on GitHub: processing of long-read data (https://github.com/holstegelab/snakemake_pipeline); and support for structural variant calling (https://github.com/holstegelab/otter; https://github.com/holstegelab/haplr).

## ACKNOWLEDGEMENTS

The authors are grateful to all study participants, their family members, and the participating medical staff involved in patient diagnosis, blood collection, blood biobanking, DNA preparation and sequencing.

Part of the work in this manuscript was carried out on the Dutch national e-infrastructure with the support of SURF Cooperative. Computing hours were granted to H. H. by the Dutch Research Council (‘100plus’: project# vuh15226, 15318, 17232, and 2020.030; ‘Role of VNTRs in AD’; project# 2022.028, 2024.036, ‘Alzheimer’s Genetics Hub’ project# 2022.31, 2025.046). This work is supported by a VIDI grant from the Dutch Scientific Counsel (#NWO 09150172010083) and a public-private partnership with TU Delft and PacBio, receiving funding from ZonMW and Health∼Holland, Topsector Life Sciences & Health (PPP-allowance), and by Alzheimer Nederland WE.03-2018-07 and WE.03-2025-08. H.H. and S.v.d.L. are recipients of ABOARD, a public-private partnership receiving funding from ZonMW (#73305095007) and Health∼Holland, Topsector Life Sciences & Health (PPP-allowance; #LSHM20106). S.v.d.L. is a recipient of ZonMW funding (#733050512). H.H. was supported by the Hans und Ilse Breuer Stiftung (2020), Dioraphte 16020404 (2014) and the HorstingStuit Foundation (2018). Acquisition of the PacBio Sequel II long-read sequencing machine was supported by the ADORE Foundation (2022).

## AUTHOR CONTRIBUTIONS

H.H. and A.N.S. conceived and designed the study. A.N.S. and N.T. curated the genetic data, with S.v.d.L. providing expertise in genetic analysis. L.K., S.W. and J.K. contributed to the generation of the long-read sequencing data. M. Huisman and N.M.v.S. contributed data from the LASA cohort, which was used as an age-matched non-demented control cohort. B.T., J.V., and M.R. contributed to the Amsterdam Dementia Cohort, which was used as the AD-case cohort. M. Hiltunen, M.D., L.P., A.A.D. and I.R.H. provided expertise in *TMEM106B* biology and related proteinopathies. A.N.S. performed the statistical and computational analyses and interpreted the results, with M. Hulsman providing computational expertise. A.N.S. and H.H. drafted the manuscript. All authors reviewed and approved the final manuscript. H.H. and S.v.d.L. supervised the study and acquired funding.

