## Supplementary Information for "Fine-mapping reveals four *TMEM106B* haplotypes associated with neurodegenerative risk and preserved cognition at extreme age"

### SUPPLEMENTARY RESULTS

#### ***Haplotype assignments from haplr are highly concordant with Hifiasm and HiPhase***

We evaluated locus- and SV-level haplotype assignments from haplr by comparison with whole-genome *de novo* assembly using *hifiasm* and reference-based long-read phasing using HiPhase **(Supplementary Methods)**. Across 493 samples, corresponding to 986 haplotype copies, *hifiasm* completely resolved both haplotypes across the *TMEM106B* locus in 338 samples (69%; 676 of 986 haplotype copies). *HiPhase* completely resolved the locus in 407 samples (83%; 814 of 986 haplotype copies), whereas *haplr* provided complete haplotype assignments in 492 samples (99.8%; 984 of 986 haplotype copies).

Among the 338 samples with complete results from both *haplr* and *hifiasm*, diplotype assignments were concordant in 337 samples (99.7%). Similarly, among the 407 samples with complete results from both *haplr* and *HiPhase*, diplotype assignments were concordant in 406 samples (99.8%). The discordant *hifiasm* and *HiPhase* assignments occurred in two different samples. In both cases, *haplr* assigned a T1/T2 diplotype, whereas the alternative method assigned T1/T1. Inspection of the corresponding *DeepVariant* calls and manual review of the long-read alignments in IGV (v2.19.7) supported a heterozygous genotype at the T2 marker variant and, therefore, the *haplr* assignment. Both samples had low sequencing coverage (≤9x), suggesting that limited read support contributed to the homozygous assignments made by *hifiasm* or *HiPhase*.

We next assessed SV-to-haplotype concordance for two hallmark SVs at the locus: the AluYb8 insertion and the 19-kb genomic rearrangement (**Supplementary Table S4**). Among sample-haplotype pairs for which the AluYb8 insertion was phased by both *haplr* and *hifiasm*, assignments were concordant for 259 of 269 pairs (96%). For the 19-kb rearrangement, assignments were concordant for 62 of 67 pairs (93%). Comparison of *haplr* with *HiPhase* showed concordant AluYb8 assignments for 326 of 351 overlapping sample-haplotype pairs (93%) and concordant rearrangement assignments for all 88 overlapping pairs (100%).

Consistent with these sample-level comparisons, the population-level distributions of both SVs across the T1-T4 haplotypes were similar among methods. *Haplr* assigned the AluYb8 insertion predominantly to T1 and T4, with frequencies of 99%, 0%, 0% and 89% among callable T1, T2, T3 and T4 haplotype copies, respectively. The corresponding frequencies were 97%, 0%, 0% and 80% using *hifiasm*. *HiPhase* also assigned the insertion predominantly to T1 and T4, although it more frequently assigned the insertion to T2 and T3, with frequencies of 99%, 9.4%, 6.3% and 100%, respectively.

The 19-kb rearrangement was assigned predominantly to T3 and T4 by all three approaches. The corresponding frequencies among T1, T2, T3 and T4 haplotypes were 1.0%, 1.0%, 92% and 89% using *haplr*; 1.0%, 1.5%, 100% and 100% using *hifiasm*; and 1.0%, 1.0%, 92% and 89% using *HiPhase*.

Together, these analyses show that *haplr* recapitulated the diplotype assignments and principal SV-to-haplotype relationships obtained using *hifiasm* and *HiPhase*. The main population-level conclusions were therefore robust to the choice of phasing approach, while *haplr* retained substantially more samples, particularly those with lower sequencing coverage, for haplotype-resolved analysis.

***Distinct TMEM106B haplotypes in genomes of African ancestry***

Previous studies have reported differences in linkage disequilibrium (LD) structure at the *TMEM106B* locus between individuals of European and African ancestry, suggesting that haplotypes with distinct allele combinations may occur across ancestry groups (**see Main text**).

However, the specific combinations of coding and structural variants underlying these differences remain incompletely characterized.

To investigate *TMEM106B* haplotype structure across ancestries, we compared 493 long-read Dutch genomes of European ancestry with 23 phased, long-read-based de novo assemblies from individuals of African ancestry generated by the Human Pangenome Research Consortium (HPRC). Among the Dutch genomes, 338 had two fully phased *TMEM106B* contigs assembled with *hifiasm*. All 23 HPRC genomes had two phased contigs spanning the locus; however, one contig from NA18906 showed two alternative alignments around the rearrangement breakpoint and was excluded from the primary comparison, leaving 22 HPRC genomes.

In the 338 Dutch, we confirmed strong LD between (i) the Block 2 marker SNP rs13237415 and the ~19-kbp rearrangement, and (ii) rs3173615 (Thr185Ser), the AluYb8 insertion, and rs1990620 (**Figure 3a**). In contrast, LD between these variant sets was substantially reduced in the 22 HPRC genomes (**Figure 3a**), consistent with greater recombination between alleles that are tightly linked in the Dutch genomes.

We next compared the phased *TMEM106B* assemblies from African-ancestry genomes with the T1-T4 haplotypes defined in this study **(Figure 1)**. Based on the marker variants listed in **Supplementary Table S1**, we identified three haplotypes with distinct combinations of coding and structural alleles: two T1-like haplotypes and one T3-like haplotype **(Figure 3b,c).** Notably, the T1-like haplotype carried the risk-associated 185-threonine allele while lacking the AluYb8 insertion and carrying the rs1990620-G allele, both of which have been associated with a lower risk for neurodegenerative traits. The T1-like and T3-like haplotypes also differ in the presence or absence of the ~19-kbp rearrangement **(Figure 3c)**. Functional studies will be required to determine the biological consequences of these allele combinations.

We separately evaluated the excluded HPRC genome, NA18906. Manual inspection of read alignments with samtools (v1.13) and aligned *de novo* assemblies in IGV (v2.19.7) supported homozygosity for the reference alleles at rs5011439 and rs13237415 together with the alternative allele at rs1990620, consistent with the distinct T1-like haplotype observed above. Read alignments and assemblies also supported homozygosity for the ~19-kbp rearrangement and absence of the AluYb8 insertion. These calls were independently supported by the *haplr svg* command, which infers the most likely genotype from read support (see below). Thus, NA18906 carries a distinct combination of alleles that was also observed in other genomes of African ancestry **(Figure 3c)**.

Together, these findings indicate that *TMEM106B* haplotype architecture differs between the ancestry groups examined and that variants tightly linked in the Dutch genomes occur in additional combinations in genomes of African ancestry. These haplotypes provide a framework for investigating the functional and disease-associated effects of individual *TMEM106B* variants.

### SUPPLEMENTARY METHODS

#### *PacBio whole-genome long-read sequencing*

Genomic DNA of AD and centenarian individuals were extracted from peripheral blood, and gDNA integrity was assessed with Qubit system 2 (Qubit dsDNA HS Assay Ref. No. Q32854) and Nanodrop (NanoDrop 2000). Specifically, we required DNA to have a purity of 260/230 of 2.00 and above, and 260/280 of 1.8 and above. When requirements were not met, a clean-up step was performed with 20 minutes incubation at room temperature with 1:1 AMPure PB beads (Pacific Biosciences Ref. No. 100-265-900) and two times washing with 80% ethanol. DNA is eluted with 120 µl Elution buffer (Pacific Biosciences Ref. No. 101-633-500). gDNA was sheared with the Megaruptor from Diagenode with a long hydropore cartridge (Cat. No. E07010003), using insert concentration of 116 µg with a shearing speed of first 30, and afterwards 31. Qubit dsDNA HS Assay (Ref. No. Q32854) was used to quantify DNA concentration. DNA damage repair was performed using SMRTbell Enzyme Clean Up Kit 2.0 (Ref. No. 101-932-600) with the reaction volumes as described in the manufacturer's instructions. More specifically, a nuclease treatment on the formed SMRTbell library was done by preparing a master mix with SMRTbell Enzyme Clean Up Mix (Pacific Biosciences Ref. No.101-932-900), SMRTbell Enzyme Clean Up Buffer 2.0 (Pacific Biosciences Ref. No. 101-932-700) and Molecular biology Grade water. Reaction mix was added to the library complex and incubated at 37°C for 30 minutes. SMRTbell Library was purified using 1.x AMPurePB beads (Pacific Biosciences Ref. No. 100-265-900) and two times washing with 80% ethanol, library complexes were eluted in 31 µl Elution buffer (Pacific Biosciences Ref. No. 101-633-500).

All libraries were prepared using either SMRTbell Template Prep Kit 2.0 (Pacific Biosciences Ref. No. 100-938-900) or 3.0 (Ref. no. 102-141-70) following manufacturer’s instructions. Size selection was performed using the BluePippin high-pass DNA size selection (sage Science

BLU0001; 0.75% Agarose Cassettes, with Marker S1). The range selection mode was set from 8,000-50,000 bp. Qubit dsDNA HS assay was used to quantify DNA concentration. Libraries having the desired size distributions were identified on the Femto pulse System (Agilent M5330AA) using the 48kb Ladder. Fractions centered between 15 kb, and 21 kb were used for sequencing.

Polymerase binding for all sequencing libraries were prepared using Sequel® II Binding Kit 2.2 (Pacific Biosciences Ref. No. 101-894-200) and 3.2 (Pacific Biosciences Ref. No. 102-194-100) following manufacturer's instructions. Sequencing primers were conditioned by heating to 80°C for 2 minutes and rapidly cooled to 4°C. The sequencing primer was annealed to the template at a molar ratio calculated by PacBio for 15 minutes at room temperature. After primer annealing, polymerase was bound to the primed template for 15 minutes at room temperature. Polymerase-bound samples were then kept at 4°C prior to use. Prior to sequencing, excess unbound polymerase was removed by incubating the complexes for 5 minutes with 1.2× (vol:vol) AMPure PB beads (Pacific Biosciences Ref. No. 100-265-900) at room temperature. Beads were not washed with 80% ethanol. After removing the free polymerase in the supernatant, the polymerase-bound complexes were eluted with Adaptive loading buffer (Pacific Biosciences Ref. No. 102-030-300). Sequencing reaction was performed using the Sequel sequencing kit 2.0 (ref. no. 101-820-200) on a SMRT-cell 8M chip (ref. no. 101-389-001).

All samples were sequenced at the Department of Clinical Genetics at the Amsterdam University Medical Center (AUMC) using a Pacific Biosciences (PacBio) Sequel IIe System with 2 hours of pre-extension time and 30 hours of collection time. Overall, 92 AD genomes were sequenced at a median coverage of 18.2x, with median read-lengths of 14.8 Kbp; 117 centenarian genomes were sequenced at a median coverage of 20.1x, with median read-lengths of 14.7 Kbp.

***PacBio long-read sequencing data processing***

After sequencing, raw reads were collected and analyzed through an in-house pipeline (freely available at https://github.com/holstegelab/snakemake_pipeline). Briefly, raw reads first underwent PacBio's ccs algorithm (v6.0.0, https://github.com/PacificBiosciences/ccs) to generate high-fidelity (HiFi) reads with custom parameters (min-passes 0, min-rq 0, keeping kinetics information). Using these parameters, we retained both HiFi reads (read-quality >99%, number of passes >3), and lower-quality non-HiFi reads, which are typically excluded when running the ccs algorithm with default settings. HiFi and non-HiFi reads were then aligned to GRCh38 (patch release 14) reference genomes using pbmm2 (v1.3.0, https://github.com/PacificBiosciences/pbmm2), with suggested alignment presets for HiFi and non-HiFi reads, generating one BAM-file per SMRT-cell. When available, individual BAM-files from the same sample were merged into a single BAM-file using *samtools* (v1.13).

***In-house long-read SV-calling pipeline, phasing, and haplotype-block matrix***

Candidate SVs in the *TMEM106B* locus were first identified using sniffles2 (version 2.0.6) with default parameters, and their respective coordinates were converted to a BED-file. Tandem repeat (TR) annotations were then downloaded from the UCSC genome browser (https://genome.ucsc.edu/), and their coordinates were merged with the location of the above candidate SVs, forming a list of regions representing candidate SVs. Local assembly of each region per genome was then performed with otter (v1.0) using the 'assemble' with '-x 2,0.95' parameters to control for highly erroneous singleton reads.

We developed a joint-genotyping procedure to compare and associate SVs across a large collection of genomes. This procedure is implemented in otter (v1.0) under the 'genotype' command. In short, we used otter to store locally assembled allele sequences in BAM-format, one per genome. All BAM-files are then merged into a single BAM file, effectively creating a compressed and queryable database of all allele sequences of SVs in a collection of genomes.

For each SV-region, we cluster all similar allele-sequences, and consider each cluster a unique genotype of an SV. More specifically, we represent all allele sequences by both their total length, and by a n-dimensional vector of normalised k-mer frequencies, where n represents the total possible number k-mers. We then compute two separate pairwise distance matrices for all allele sequences for each SV-region: one based on the absolute difference between allele-lengths; and another based sequence content, by computing pairwise cosine distances between the normalised vectors of the k-mer usage of each allele-sequence. We perform average-linkage hierarchical-clustering for each distance matrix, and assign cluster membership by cutting the dendrogram at a maximum input cluster distance. Two allele sequences are considered to be similar if their length and k-mer cluster-memberships are identical. Similar allele-sequences are grouped together, and the medoid of each group based on allele-length is selected as the representative allele-sequence, and hence, the corresponding genotype.

The output of the above procedure is gVCF with genotypes for each genome for each SV. Furthermore, we provide additional metadata, such as coverage support and sequence complexity, based on the Hill-Shannon diversity index of the normalised k-mer usage vector, which highlights whether an allele-sequence is highly repetitive (e.g. low diversity) or more complex (e.g. high diversity)

We showcase the joint-genotyping procedure implemented in otter, by locally assembling 1,042,912 TRs annotated across the GRCh38 reference genome, and genotyping the allele-sequences of each TR across the 513 AD and centenarian genomes. The TR-annotations were compiled from TR-annotations from the UCSC Genome Browser, PacBio TR catalogue, and the ADOTTO TR catalogue. Using 10 threads on a compute node with an AMD Rome 64 processor and local SSD storage, we were able to genotype the alleles of all TRs from all genomes in 34 mins, with a maximum RAM usage (RSS) of 0.41 Gbs.

Finally, to ensure high sensitivity and accuracy for genotyping the genomic rearrangement, we chose a known high-coverage T1 and T3 heterozygous carrier based on array imputation, and performed *de novo* assembly with *hifiasm* (v0.16.1-r375), and aligned the resulting phased contigs to the GRCh38 genome. We then genotyped the local sequence content of rearrangement breakpoint (chr7:12160056-12160400) with the 'wgat' command in *haplr* (v0.1.0), generating representative allele sequence(s) per genomic rearrangement per SV. We used *haplr* to probabilistically infer the most likely genotype for each of the 493 long-read–sequenced genomes, together with a corresponding genotype quality (GQ) score.

***Haplotype-guided assignment of long reads to predefined GWAS haplotypes using Haplr***

Long-read sequencing enables genetic and epigenetic variation to be resolved on individual DNA molecules across extended genomic regions. Existing read-based phasing tools, including WhatsHap and HiPhase, reconstruct sample-specific haplotypes by grouping reads according to heterozygous variants identified in the sequencing data. De novo assembly approaches such as Hifiasm instead reconstruct haplotypes without requiring heterozygous variants to be specified a priori. Both strategies can support haplotype-resolved analyses, although additional processing is generally required to quantify haplotype-specific methylation. Their performance may also be limited at low sequencing coverage. Among 493 long-read-sequenced genomes, the *TMEM106B* region could be completely assembled and phased end to end in 338 samples (69%) using hifiasm and completely phased in 404 samples (82%) using HiPhase, with incomplete phasing occurring predominantly in samples with less than 10× coverage.

We therefore sought to use prior haplotype information to characterize structural variation and DNA methylation in samples that could not be completely phased using these approaches. At many GWAS loci, the combinations of SNPs and small insertions and deletions that define common disease-associated haplotypes can already be inferred accurately from array-based genotype imputation. In this setting, the principal unresolved features are not necessarily the small-variant haplotypes themselves, but the structural variant alleles and methylation profiles carried on those haplotypes. Because phased, imputed haplotypes were available for all 493 long-read-sequenced samples, individual long reads could be evaluated according to their support for each predefined haplotype.

We therefore developed *haplr*, a haplotype-guided framework that probabilistically assigns individual long reads to predefined haplotypes using phased haplotype information. Rather than reconstructing haplotypes *de novo*, *haplr* leverages existing GWAS haplotype definitions to enable haplotype-resolved analysis of structural variants and CpG methylation, including in samples with lower sequencing coverage. This approach provides a direct link between established GWAS haplotypes and molecular features observed in long-read sequencing data.

Briefly, *haplr* takes aligned long reads in BAM format together with predefined haplotypes specified using single marker variants, multivariant haplotype definitions or nested sub-haplotypes derived from phased GWAS or reference-panel data. Rather than reconstructing haplotypes *de novo*, *haplr* evaluates the support of each read for each candidate haplotype using a probabilistic model that incorporates allele concordance, sequencing error probabilities and, optionally, haplotype-frequency priors. Each read is assigned to the most likely haplotype and reported with a posterior probability reflecting assignment confidence. *Haplr* additionally supports haplotype-resolved SV genotyping and genotype recalibration, as well as CpG methylation profiling. Full model formulations and implementation details are provided in the Supplementary Note, "Probabilistic model and implementation of *haplr*." *Haplr* is implemented in C++ as a standalone command-line tool and publicly available through github (https://github.com/holstegelab/haplr).

### ***Phasing benchmark between haplr with hifiasm and Hiphase***

We benchmarked *haplr* by comparing the inferred haplotype and SV-to-haplotype phasing statistics across our 493 samples with that of *hifiasm* (whole-genome *de novo* assembly) and *HiPhase* (reference-based long-read phasing). Briefly, we first generated complete *de novo* assemblies with *hifiasm* (v0.16.1-r375) using default parameters, and aligned the resulting phased-contigs to GRCh38 reference using *minimap2* (2.21-r1071) using the '-ax asm10' parameter; the alignments were stored as individual BAM files. We then used *haplr*'s *wgat* command to extract all contig-alignments at the *TMEM106B* locus, and extract the corresponding allele sequence at each expected SV location based on the joint SV-genotyping gVCF of the 493 samples **(see Methods)**. For each extracted allele sequence, we compared it to all possible alleles in the gVCF, and assign an allele index to each contig based on the minimum edit-distance using the edlib library (v.1.27) (10.1093/bioinformatics/btw753). For *HiPhase* (v1.4.2-c7e0700), we phased all genetic variants by providing both the joint gVCF from *DeepVariant* (v1.1.0) and *GLnexus* (v1.4.1-0-g68e25e5), and the joint gVCF from otter (v1.0), using default parameters. This approach allowed us to generated a phased and harmonized VCF per sample for both small and large (structural) variants, allowing us to systematically evaluate which alleles are present in each phased contig at the *TMEM106B* locus.

We benchmarked the three approaches using three complementary analyses. First, we quantified the number of samples for which both haplotype copies across the *TMEM106B* locus were completely resolved and could be assigned to the T1-T4 haplotypes using the marker SNPs listed in **Supplementary Table 1**. Second, among samples with complete results from both methods being compared, we assessed diplotype concordance between *haplr* and *hifiasm* or *HiPhase*. Diplotype assignments were considered concordant only when the inferred diplotype matched exactly, for example T1/T1, T1/T2 or T1/T3. Third, we assessed SV-to-haplotype phasing concordance for two hallmark SVs at the locus: the AluYb8 insertion and the 19-kb genomic rearrangement. An overlapping sample-haplotype pair was defined as a haplotype copy for which the SV was callable and phased by both methods being compared. SV-to-haplotype assignments were considered concordant only when both the SV allele and its assigned T1-T4 haplotype matched exactly. We additionally compared the population-level frequencies of each SV across the T1-T4 haplotypes for all three approaches.

### SUPPLEMENTARY NOTE

#### ***Probabilistic model and implementation of haplr***

*1. Bayesian haplotype assignment of long-reads*

*Haplr* assigns long-read sequences to predefined haplotypes using a probabilistic framework: it treats haplotypes as latent classes, while read-level sequence observations provide evidence for assignment to a haplotype. For each primary read, *haplr* computes the posterior probability of membership in each haplotype, and reports the most likely assignment.

Given a set of predefined haplotypes at a locus, haplr models the haplotype assignment of a read using a Bayesian framework, in which the haplotype label takes values $h \in H$, where $H$ denotes the set of haplotypes under consideration. For each read, the posterior probability of originating from haplotype $h$ is proportional to the product of a haplotype prior and a read likelihood,

$$P(H = h \mid read) \propto P(H = h)P(read \mid H = h).$$

Haplotype priors are uniform by default but can be extended to reflect known haplotype frequencies. The read likelihood is computed across haplotype-informative variant sites covered by the read,

$$P(read \mid H = h) \approx \prod_{i \in I} P(b_i \mid a_{ih}, Q_i),$$

where $b_i$ and $Q_i$ denote the observed base and base quality at site $i$, respectively, and $a_{ih}$ is the allele carried by haplotype $h$ at that position.

At each informative position (positions that are covered by a read), *haplr* compares the observed allele to the expected allele for each haplotype. To account for sequencing errors, base quality scores are converted to error probabilities using standard phred scaling under the following substitution model,

$$P(\, b_i \mid a_{ih}, Q_i \,) = \begin{cases} 1-\varepsilon_i & match \\ \varepsilon_i/3 & mismatch \end{cases},$$

where $\varepsilon_i$ denotes the base-calling error probability derived from the effective phred quality score at site $i$. Sites failing minimum quality thresholds or carrying undefined haplotype alleles are ignored, effectively marginalizing missing observations.

*Haplr* then sums the log-likelihoods across informative positions to calculate a haplotype-specific score corresponding to the log joint probability,

$$score_h = \; log(P(H=h)\prod_{i \in I} P(b_i \mid a_{ih}, Q_i)).$$

Following Bayes' rule, the posterior probability follows:

$$P(H=h \,|\, read) = \frac{\mathrm{e}^{\mathrm{score}_h}}{\sum_{k=1}^{n} \mathrm{e}^{\mathrm{score}_k}} \,.$$

Reads are assigned to the haplotype with the maximum posterior probability. To account for uncertainty arising from limited evidence, haplr requires a minimum number of informative sites and a user-defined posterior confidence threshold; reads failing these criteria remain unassigned.

To support sub-haplotype assignment, *haplr* allows multiple haplotype label sets (e.g. main haplotypes and nested sub-haplotypes) to be evaluated independently. For sub-haplotype assignment, *haplr* first restricts the analysis to positions that are informative with respect to the corresponding main haplotype background, excluding sites that do not distinguish the sub-haplotypes within that background. Haplotype assignment is then performed as a separate probabilistic competition using the same read evidence. This design allows sub-haplotype assignments to remain informative even when main haplotype assignments are ambiguous, such as when sub-haplotypes are defined by a small number of highly informative variants.

*2. Likelihood assignment of known structural variants*

Given a known structural variant (SV; i.e. from a joint-VCF file) with a set of alleles, $A = \{a_1, a_2, \dots, a_m\}$, haplr will extract all primary read-sequences, $S = \{s_1, s_2, \dots, s_r\}$, and calculate the log-likelihood of each sequence originating from one of the SV-alleles,

$$\Lambda_{r,m} = \log\left(P(s_r \mid a_m)\right) .$$

For a given genotype, $G = (i, j) \mid 1 \le i \le j \le m$, we calculate the probability of observing $s_r$ given $G$,

$$P(s_r \mid G = (i,j)) = \log\left(0.5 e^{\Lambda_{s_r,i}} + 0.5 e^{\Lambda_{s_r,j}}\right).$$

And the log-likelihood that the read-set $S$ originates from a given $G$,

$$\log\left(P(S \mid G)\right) = \sum_r \log\left(P(s_r \mid G)\right)$$

The genotype that maximizes the log likelihood, $\hat{G}$, is selected as the final genotype of the sample,

$$\hat{G} = \arg\max \log\left(P(S \mid G)\right) .$$

We detail this likelihood framework below.

*2.1 Computing a likelihood based on sequence similarity and spanning-context*

For simplicity, we refer to sequence $s_r$ as $r$, and allele $a_m$ as $m$. We define $M_{r,m}$ as the joint likelihood that $r$ originates from $m$ based on sequence similarity $S_{r,m}$, and spanning-context, $L_{r,m}$:

$$M_{r,m} = S_{r,m} + L_{r,m} \ .$$

We consider three situations for a given read-alignment: (a) it fully overlaps the reference coordinates of a SV; (b) partial overlaps (only left, right, or contained); (c) the read does not overlap and hence contains no sequence information. In implementation, we skip read-alignments from situation (c).

To calculate $S_{r,m}$, we first compute the edit distance between $r$ and $m$, referred to as $d_{r,m}$. We account for the spanning context of $r$: Needleman-Wunsch alignment (situation a); Smith-Waterman (situation b left/right spanning); or Hirschberg's algorithm (situation b contained). We set the total sequence length as $N_{r,m} = \max(|r|, |m|)$. For an expected sequencing error rate, $\varepsilon$, we calculate the likelihood of sequence $r$ originates from $m$ based on sequence similarity,

$$S_{r,m} = d_{r,m} \log(\varepsilon) + \left(N_{r,m} - d_{r,m}\right) \log(1 - \varepsilon) \ .$$

The length of sequence $r$ provides important information. Consider that two possible alleles exist, one with 10 bps and another at 100 bps. A read-sequence of 50 bps provides important clues about the origin of the sequence: if $r$ is fully spanning, then it might be a highly erroneous sequence, and/or a sequence not originating from the current genomic locus of interest (i.e. misalignment). But if $r$ only partially spans or is fully contained, then we must consider the possibility that it may originate from the allele of 100 bps instead of 10 bps. This is contrast to a sequence that is 5 bps, which (based on length alone) is ambiguous as it may originate from either allele.

The $L_{r,m}$ tries to assess whether $|r|$ given $|m|$. First, we set two parameters: $\gamma_m$ specifies the number of additional bases $r$ is 'allowed' to have relative to $|m|$ before penalizing it. It is defined by $\gamma_m = \lceil c_\gamma \varepsilon |m| \rceil$, where $c_\gamma$ is a user-defined penalty factor and $\varepsilon$ is the expected sequencing error-rate. The penalty score, $\rho$, calculates the per-base penalty when $r$ is 'too large', scaled to the expected error-rate and user-defined penalty-factor $\rho = (-c_p \log(1-p))$.

If $r$ is fully spanning, we define,

$$L_{r,m} = -\rho\big(\big||r| - |m|\big| - \gamma_m\big), \text{ if } \big||r| - |m|\big| > \gamma_m, \text{ and 0 otherwise.}$$

If $r$ is partially spanning or contained, we set $L_{r,m}$ as the joint likelihood that $|r|$ may plausibly originate from allele $m$ based on $|m|$, defined as $L_{r,m}^{feas}$; and whether $|r|$ is informative (i.e. unambiguous) given $|m|$, $L_{r,m}^{info}$:

$$L_{r,m} = L_{r,m}^{feas} + L_{r,m}^{info}$$

$$L_{r,m}^{feas} = -\rho \max\big(0, |r| - (|m| + \gamma_m)\big)$$

$$L_{r,m}^{info} = \alpha \log\left(\frac{\min(L,A)+1)}{A+1}\right).$$

Here, $\alpha$ is a user-defined parameter that controls the strength of $L_{r,m}^{info}$, where $\alpha = 0$ disables this term, and $\alpha = 1$ scales it linearly, and $\alpha > 1$ strengthens the term.

*2.2 A mixture model to account for highly erroneous read-sequences*

Finally, we consider that ONT and PacBio can generate (singleton) highly erroneous read-sequences, such as those from low-complexity regions. To prevent these outlier sequences from dominating the likelihoods across a given set of alleles, we define two user-provided parameters: (i) $\pi$, the probability of observing an outlier sequence; and (ii) $\beta$, which is a factor that defines a maximum penalty for sequence $m$, such that $B_{r,m} = \beta N_{r,m}$.

We thus 'cap' the final log-likelihood that $r$ originates from $m$ by accounting for such possible outlier sequences:

$$\Lambda_{r,m} = \log\left(P(s_r \mid a_m)\right) = \log\left((1-\pi)e^{M_{r,m}} + \pi e^{B_{r,m}}\right)$$

*3. Structural variant calling and methylation profiles*

To support downstream analyses, haplr can generate haplotype-specific methylation profiles and structural variant (SV) detection based on per-read haplotype assignments. Methylation calls reported in the input BAM file are aggregated by assigned haplotype to estimate per-CpG methylation fractions, accounting for potential reference allelic biases, and subject to a user-defined minimum methylation probability threshold.

For SV detection, haplr uses a user-provided joint VCF that defines candidate SVs. For each candidate SV, haplr evaluates read-level support by locally aligning reads to each possible SV allele, and aggregating the alignments under the specified likelihood model above to infer the most likely SV-genotype, which is reported together with a genotype confidence score, GQ. GQ is calculated as follows: a delta is calculated for the top two genotype log-likelihoods, $\Delta = G_{best} - G_{second}$. These are then converted to a Phred-scaled likelihood represented as an integer capped at 99, similarly to GATK: $GQ = \left\lfloor 10log_{10}\left(1 - e^{\Delta}\right) + \frac{1}{2} \right\rfloor$.

*4. Implementation details*

*Haplr* is implemented in C++ and makes use of the *HTSLIB* library (v1.23) for accessing and processing VCF and BAM files, and *edlib* library (10.1093/bioinformatics/btw753) to calculate alignments and edit-distances.
